# Reducing Biosecurity Risks in Informal Poultry Slaughter: A Pilot Intervention Among Mobile Poultry Vendors in Bangladesh

**DOI:** 10.64898/2026.11.05.26367345

**Authors:** Md. Musa Baker, Kamal Ibne Amin Chowdhury, Dalia Yeasmin, Abu-Hena Mostofa Kamal, Mohammad Tauhidul Islam, Sheikh Shariful Islam, Ausraful Islam, Md. Saiful Islam

**Affiliations:** International Centre for Diarrhoeal Disease Research, Bangladesh (icddr,b), Dhaka-1212, Bangladesh; Khulna University of Engineering & Technology (KUET), Bangladesh; School of Population Health, University of New South Wales, Sydney, Australia

**Keywords:** Avian Influenza, poultry vendors, safe slaughtering, waste disposal, Bangladesh

## Abstract

**Introduction:** Mobile poultry vending and slaughtering practices in Bangladesh pose potential public health risks due to poor hygiene, unsafe waste disposal, and close human-animal interactions. Limited data exist on these practices and feasible interventions to improve biosecurity among mobile poultry vendors.

**Methods:** This mixed-method study was conducted in three phases across four sub-districts in three districts. Phase 1 involved 416 hours of structured observation, 40 in-depth vendor interviews, and 40 informal customer interviews. Phase 2 involved the development and pilot testing of an intervention package, including poultry killing cones, hand sanitizers, disinfectants, and a hygiene pamphlet, with 10 vendors. Phase 3 implemented the intervention among 20 vendors for one month, followed by 94 hours of observation and 17 customer interviews.

**Result:** Phase 1 found 1,959 slaughtering events with no use of personal protective equipment; slaughtering occurred in plastic jars (58%), directly on roads (21%), and roadside drains (11%). Seventy percent of vendors reported discarding slaughtering waste directly into the environment, while 78% reported not knowing how poultry diseases could spread to other animals or humans. During Phase 2, 9/10 vendors considered the killing cone useful, although bicycle users reported challenges related to weight and balance; the cone was subsequently modified. During Phase 3, 294 poultry were observed being slaughtered, with 79% using killing cones. Waste was stored in cone-attached boxes in 97% of cone uses, and vendors used hand sanitizer before eating in 75% of observed instances. Among 17 interviewed customers, 88% recognized the cone’s role in safer waste disposal and 53% considered it useful overall.

**Conclusion:** This study highlights major hygiene and biosecurity gaps and demonstrates that low-cost, targeted interventions may improve biosecurity practices among mobile poultry vendors and reduce opportunities for environmental contamination in informal slaughter settings.

## Introduction

In Bangladesh, Avian Influenza A (H5N1) virus is endemic in poultry (1). Since 2007, nine human cases of H5N1 have been reported, including one death (2). All cases had a history of exposure to poultry or poultry products and waste before illness (3, 4). For example, on March 15, 2011, urban surveillance by icddr,b in Kamalapur, Dhaka, identified a human case infected with highly pathogenic avian influenza (HPAI) H5N1 virus (4). Seven days before the onset of illness, the patient’s mother purchased seven chickens from a mobile poultry vendor. The vendor slaughtered, de-feathered, and skinned the chickens. The patient’s father further processed them inside the household, while the patient was present at the slaughtering site (4).

Mobile poultry vendors commonly sell chickens, pigeons, geese, and ducks using bicycles, a pedal-powered tricycle van, or by carrying birds by hand. They may source birds of different species from multiple locations and keep them together in close confinement during transport and sale, creating opportunities for avian influenza circulation, inter-species transmission, and viral reassortment (5–8). Vendors often slaughter poultry at the point of sale and discard blood, feathers, and other waste directly into the environment. Improper disposal of slaughter waste and carcasses into the environment is a potential source of exposure to avian influenza or other pathogens for dogs, cats, and other domestic animals (9). A community-based ethnographic study in Bangladesh reported that during poultry slaughter, humans, domestic animals, and scavenging birds are frequently exposed to poultry blood, feathers, internal organs, and other processed byproducts discarded into the open environment (10). Beyond avian influenza, mobile poultry vending may contribute to broader zoonotic and environmental health risks, including bacterial contamination and the spread of antimicrobial-resistant organisms, particularly in settings with limited sanitation and biosecurity (11, 12). Avian influenza viruses circulate naturally among wild waterfowl and can spill over to domestic poultry through shared water sources and trading networks (13, 14). In Bangladesh’s densely populated poultry sector, small-scale and backward farms dominate, while rapid urbanization has increased the informal and peri-urban poultry marketing system (10).

Surveillance databases and outbreak reports from the World Health Organization (WHO), World Organization for Animal Health (WOAH), and the Food and Agriculture Organization of the United Nations (FAO) have documented recurrent avian influenza activity in Bangladesh and across South Asia, highlighting the public health relevance of informal poultry trading and on-site slaughtering practices (15–17). Most existing research has focused on live bird markets and formal slaughterhouses; comparatively fewer studies have examined the practices, slaughter locations, and waste disposal behaviors of mobile poultry vendors, particularly in urban and peri-urban settings (10, 18). The live-bird market system in Bangladesh comprises fixed markets, backward sellers, and mobile vendors; together, these systems sustain poultry demand and employment but also create a continuum of potential exposure pathways. Mobile vendors are uniquely important because they connect rural production systems with urban consumers, often moving between communities without formal biosecurity oversight (10, 19, 20).

Despite the importance, there is limited empirical evidence on the day-to-day practices of mobile poultry vendors, including sourcing, transport, slaughtering, hygiene, and waste management behaviors. Our previous scoping work identified gaps in hygiene practices and risk awareness during on-site slaughtering and carcass disposal, highlighting the need for context-specific interventions. While mobile poultry vending is an essential component of the poultry value chain, the close interaction between humans, animals, and the environment in these settings underscores the importance of improving hygiene and reducing exposure pathways. This study aimed to assess existing poultry handling, slaughtering, and waste disposal practices among mobile poultry vendors; develop and pilot a practical, low-cost hygiene and biosecurity intervention package; and evaluate its feasibility, acceptability, and potential to improve hygiene and biosecurity practices and reduce potential exposure risks.

## Materials and methods

### Study site

The team, consisting of social scientists, medical technicians, and a veterinary epidemiologist, each with several years of experience in zoonotic disease research, conducted this study in Mirpur and Mohammadpur (sub-districts of Dhaka) and in Mymensingh Sadar and Netrokona Sadar (sub-districts outside Dhaka) (Fig. 1). The study areas were selected purposively based on the availability of mobile poultry vendors, logistical feasibility due to the presence of ongoing avian influenza surveillance in nearby live-bird markets, and the opportunity to leverage existing research infrastructure and staff. Avian influenza surveillance in nearby live-bird markets facilitated our access to both the communities and the mobile poultry vendors. Because surveillance was limited to live bird markets while this study focused on mobile poultry vendors, this introduced bias is unlikely, as mobile poultry vendors and live bird market workers operate independently, sell poultry in different ways, and at different locations.

**Fig 1.**
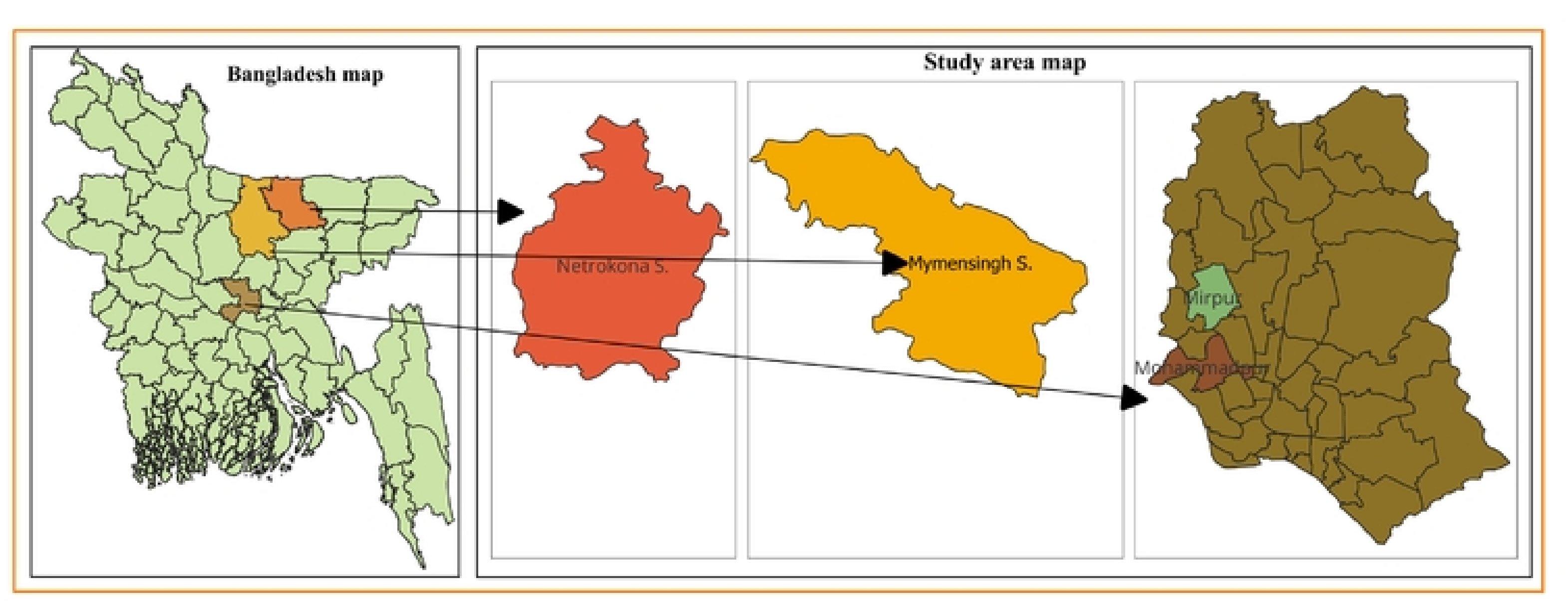
Geographic distribution of study locations across four sub-districts in Bangladesh.

### Study design and data collection

Between April 1, 2016, and March 31, 2017, we conducted this formative mixed-method study in three phases (Fig. 2). The study was guided by a grounded theory approach, allowing themes to emerge inductively from interviews and observations to explain factors influencing the acceptability of the poultry killing cone. Phase 1 included line listing of vendors, structured observation, in-depth interviews with vendors, and informal interviews with customers. A comprehensive list of mobile poultry vendors was developed in collaboration with community leaders and live bird market workers, and through snowball sampling to identify additional participants. To ensure adequacy of the sample size, we relied on data saturation. From the sampling frame, ten vendors were purposively selected from each of the four sub-districts to represent different transport types (bicycle, tricycle, and hand-carry) and diverse selling locations. We conducted 416 hours of structured observation of 40 vendors, observing each vendor from poultry collection through the end of daily sales over two consecutive days. This approach allowed us to capture variability in trading practices and to build rapport with participants, facilitating more open discussion during interviews. Trained social science researchers documented the number of poultry carried, the presence of apparently sick poultry, interactions with the customers, direct handling of poultry, slaughtering practices (who, where, and how), and disposal of blood and other waste. Poultry showing visible lethargy, ruffled feathers, nasal discharge, closed eyes, or reluctance to move were considered as apparently sick poultry during (21).

**Fig 2.**
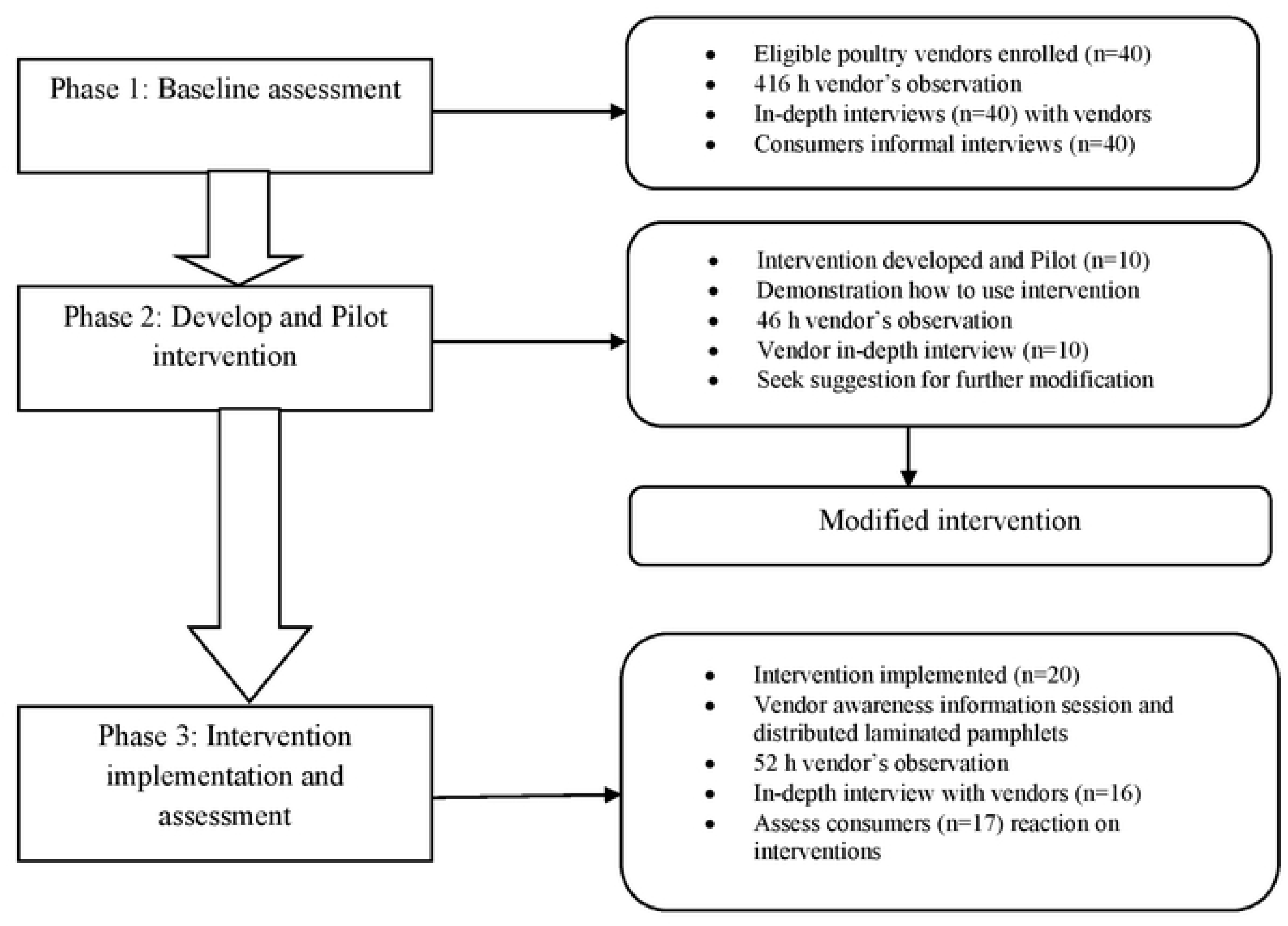
Overview of the three phases of research activities conducted in the study.

In Phase 1, we conducted 40 face-to-face in-depth interviews with the vendors observed to explore poultry trading, slaughtering, and waste disposal practices, and to clarify observation findings. Additionally, 40 informal interviews were conducted with purposively selected customers to understand purchasing behavior, slaughtering preference, waste disposal practices, knowledge on avian influenza, and suggestions for risk reduction. Customers were approached immediately after purchasing poultry, provided with study information, and interviewed upon consent using a standardized guide. The interview guides were developed collaboratively by the research team and pre-tested to ensure clarity and comprehension. Interviews were conducted in participant-preferred locations and audio-recorded with consent.

In phase 2, we developed and piloted an intervention package that included a poultry killing cone co-designed with vendors. Two cone sizes were developed to fit tricycles (Size-1: upper end 6”, lower end 1”) and bicycles (Size-2: upper end 6”, lower end 1”). The cones were complemented with a waste collection box (20” × 14” × 4”) and an underlying tray (20” × 10” × 3”) to collect blood, with sand or sawdust used for absorption. The intervention package also included hand sanitizer, soap, detergent, sodium hypochlorite, a spray bottle, and an informational pamphlet illustrating hand hygiene and poultry waste disinfection practices. Before piloting, the intervention team demonstrated proper cone use, preparation of disinfectant solutions, and handwashing practices. The intervention was pilot-tested among 10 vendors (eight using tricycle vans and two using bicycles) for five consecutive days to assess its practicality, acceptability, and usability. During this period, an independent observation team conducted unannounced structured observations to document operational practices. Although brief, the five-day pilot provided rapid, actionable feedback that informed refinement of the cone design, disinfection procedures, and training messages. Semi-structured interviews were conducted with vendors to assess feasibility and gather recommendations for design improvement. Based on the feedback, the cone design was subsequently modified before Phase 3. The upper opening was widened from 6” to 8” to accommodate larger poultry, and the frame was replaced with a lighter metallic structure to reduce weight and improve stability, particularly for bicycle users.

In phase 3, the refined intervention package was provided to 20 mobile poultry vendors for one month. Vendors were purposively selected from the phase 1 sampling frame to ensure diversity in location, vendor type, and scale of operation. Vendors who participated in the baseline and pilot phases were excluded to avoid prior exposure influencing outcomes. Each vendor received a killing cone appropriate to their transport mode (bicycle or tricycle van). In locations with temporary selling sites, shared cones were installed, and one vendor, selected by peers, was responsible for maintaining hygiene, disinfection, and waste management. All vendors were briefed on the study objectives, and participation was voluntary. To minimize the potential selection bias, all eligible vendors in the study areas were invited to participate. During the one-month implementation period, a separate trained observation team conducted 94 hours of unobtrusive observations, defined as non-participatory observation of behavior in natural settings. This approach is particularly useful for evaluating the effectiveness of environmental interventions that aim to influence health-related behavior (22). Observers were present but did not interfere with vendor activities. The team systematically recorded practices related to slaughtering, cleaning, and waste management as public behavior. In addition, we conducted 20 in-depth interviews with participating vendors and 17 informal interviews with customers who purchased poultry from these vendors. Customers were recruited using the same approach as in earlier phases: they were approached immediately after purchase, informed about the study, and interviewed upon providing written consent. These interviews explored perceptions, satisfaction, and feedback regarding improvements in slaughtering and waste management practices associated with the intervention.

### Data analysis

All quantitative data were entered into and analyzed in Microsoft Excel using descriptive statistics. For qualitative data, the research team expanded field notes and transcribed all interviews verbatim in Bengali using Microsoft Word. A preliminary coding framework was developed based on the initial research questions and study objectives, and subsequently refined through iterative reading and discussion among the investigators to incorporate emerging themes. The transcripts were then manually coded using the finalized coding framework. Coded segments were organized thematically and summarized across key domains aligned with the study objectives. Data triangulation was performed by comparing findings across different sources, including interviews, field notes, and structured observations, to enhance the credibility and depth of interpretation. We collaboratively reviewed the coding process and thematic synthesis to improve reliability and minimize individual bias.

### Ethics statement

We obtained informed written consent from all participants before conducting observations and interviews. We also sought written consent from the customers before conducting informal interviews. The study protocol was reviewed and approved by icddr,b’s institutional review board protocol # 15037.

## Results

### Phase 1: Mobile poultry trading practices

#### Socio-demographic characteristics

All vendors in Phase 1 were male and reported purchasing live poultry from wholesale live-bird markets and selling them door-to-door in residential areas. Most vendors used tricycle vans, while others relied on bicycles, carried poultry by hand, or used baskets (Table 1).

**Table 1.** Socio-demographic characteristics of the participants.

|  | Phase 1 |  | Phase 2 |  | Phase 3 |  |
|---|---|---|---|---|---|---|
| **Characteristic** | **Vendors % (n/N)** | **Customers % (n/N)** | **Vendors % (n/N)** | **Customers % (n/N)** | **Vendors % (n/N)** | **Customers % (n/N)** |
| **Age (Years)** |  |  |  |  |  |  |
| Mean | 34 | 37 | 32 | 36 | 33 | 38 |
| **Sex** |  |  |  |  |  |  |
| Male | 100 (40/40) | 30 (12/40) | 100 (10/10) | 50 (5/10) | 100 (20/20) | 53 (9/17) |
| **Marital status** |  |  |  |  |  |  |
| Married | 90 (36/40) | 100 (40/40) | 90 (9/10) | 90 (9/10) | 90 (18/20) | 88 (15/17) |
| **Religion** |  |  |  |  |  |  |
| Muslims | 100 (40/40) | 95 (38/40) | 100 (10/10) | 90 (9/10) | 100 (20/20) | 94 (16/17) |
| **Educational status** |  |  |  |  |  |  |
| No formal education | 20 (8/40) | 20 (8/40) | 30 (3/10) | 10 (1/10) | 20 (4/20) | 6 (1/17) |
| Primary education | 40 (16/40) | 40 (9/40) | 40 (4/10) | 30 (3/10) | 40 (8/20) | 29 (5/17) |
| ≥ Secondary education | 35(14/40) | 35 (11/40) | 30 (3/10) | 50 (5/10) | 35(7/20) | 47 (8/17) |
| ≥ Higher secondary | 5 (2/40) | 5 (12/40) | 0 (0/10) | 10 (1/10) | 5 (1/20) | 18 (3/17) |
| **Monthly income (BDT)** |  |  |  |  |  |  |
| Mean | 16,025±5245·20 | 29,712±28,001 | 19,000±6733·00 | 30,546±25,003 | 16,825±5245·20 | 32,765±24,098 |
| **Alternative occupation** |  |  |  |  |  |  |
| No alternative | 47 (19/40) | 90 (36/40) | 80 (8/10) | 70 (7/10) | 45 (9/20) | 70 (12/17) |
| Agriculture | 38 (15/40) | 5 (2/40) | 10 (1/10) | 20 (2/10) | 40 (8/20) | 18 (3/17) |
| Others | 15 (6/40) | 5 (2/40) | 10 (1/10) | 10 (1/10) | 15 (3/20) | 12 (2/17) |
| **Poultry business involved persons** |  |  |  |  |  |  |
| Family member | 37 (15/40) | 5 (2/40) | 40 (4/10) | 20 (2/10) | 40 (8/20) | 6 (1/17) |
| Relatives | 25 (10/40) | 5 (2/40) | 50 (5/10) | 0 (0/10) | 15 (3/20) | 6 (1/17) |
| Friends | 8 (3/40) | 0 (0/40) | 10 (1/10) | 0 (0/10) | 10 (2/20) | 0 (0/17) |
| Nobody | 30 (12/40) | 90 (36/40) | 0 (0/10) | 80 (8/10) | 35 (7/20) | 88 (15/17) |

#### Source of poultry and trading practice

Vendors reported purchasing 20-30 poultry per day, including caged farm chickens and free-range local chickens, from wholesale or village markets. In addition to poultry, three participants reported selling water birds, including cormorants, egrets, herons, and migratory birds. Unsold poultry was typically kept in cages in their living rooms or in separate cages at night. Vendors reported selling both healthy and sick poultry, often reducing the price to sell sick poultry quickly. Some reported prioritizing the sale of sick poultry and spraying water on birds to maintain a fresh appearance. During observation, 40 vendors carried a total of 2,411 poultry; 19% appeared sick, and one died during observation (Table 2).

**Table 2.**
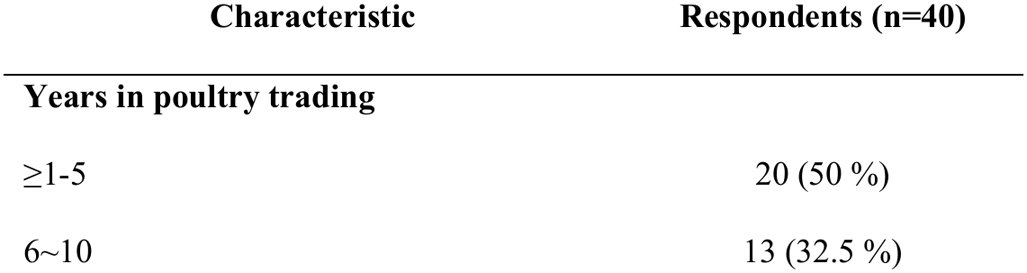
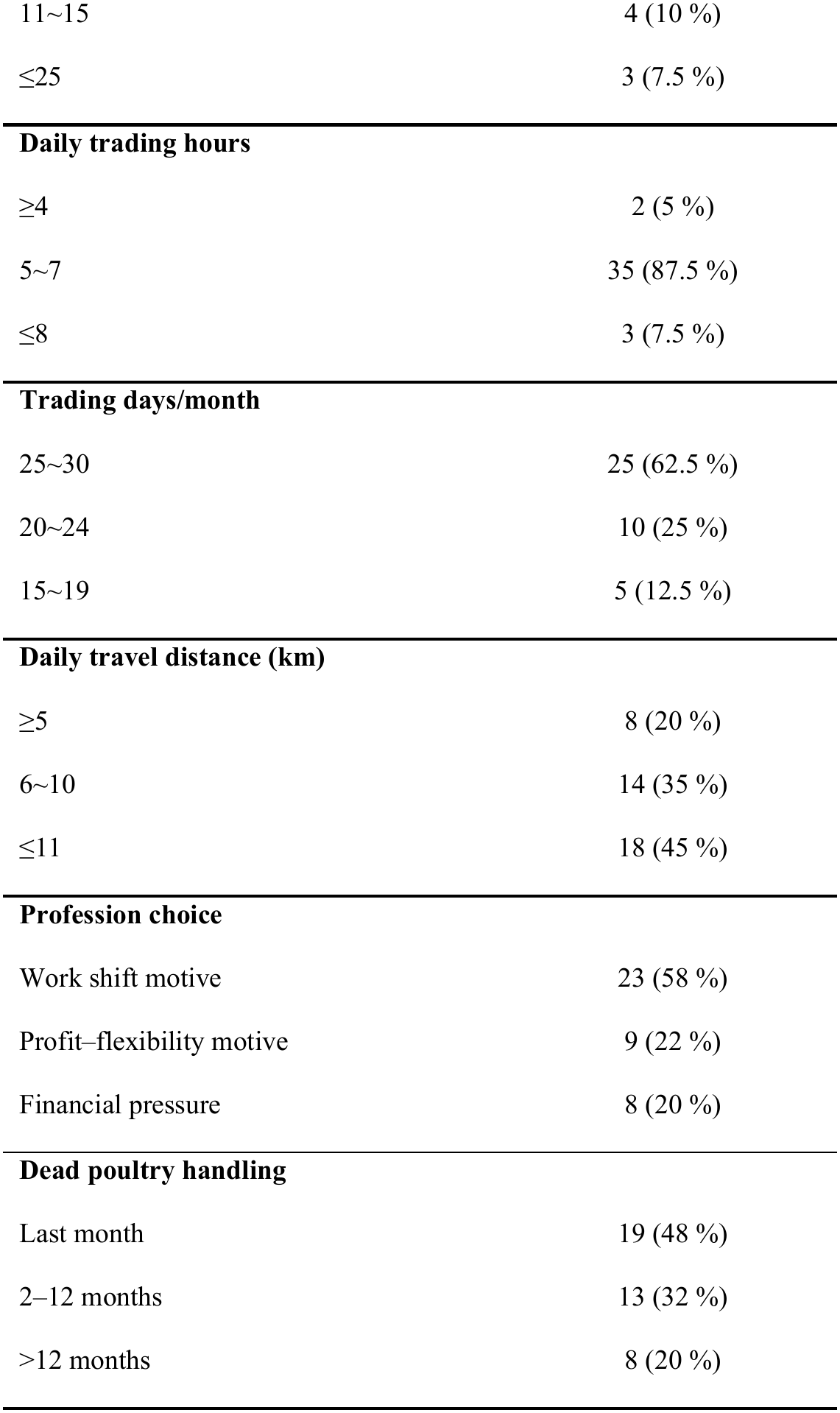
Occupational History of Mobile Poultry Vendors.

| **Characteristic** | **Respondents (n=40)** |
|---|---|
| **Years in poultry trading** |  |
| ≥1-5 | 20 (50 %) |
| 6~10 | 13 (32.5 %) |
| 11~15 | 4 (10 %) |
| ≤25 | 3 (7.5 %) |
| **Daily trading hours** |  |
| ≥4 | 2 (5 %) |
| 5~7 | 35 (87.5 %) |
| ≤8 | 3 (7.5 %) |
| **Trading days/month** |  |
| 25~30 | 25 (62.5 %) |
| 20~24 | 10 (25 %) |
| 15~19 | 5 (12.5 %) |
| **Daily travel distance (km)** |  |
| ≥5 | 8 (20 %) |
| 6~10 | 14 (35 %) |
| ≤11 | 18 (45 %) |
| **Profession choice** |  |
| Work shift motive | 23 (58 %) |
| Profit–flexibility motive | 9 (22 %) |
| Financial pressure | 8 (20 %) |
| **Dead poultry handling** |  |
| Last month | 19 (48 %) |
| 2–12 months | 13 (32 %) |
| >12 months | 8 (20 %) |

#### Poultry slaughtering and waste disposal practices

Eighty percent of vendors reported slaughtering both healthy and sick poultry. Slaughtering was commonly conducted in customers’ yards, bathrooms, inside a plastic jar, along roads, in drains, or near water sources, findings that were also confirmed through observation. We observed 1,959 slaughtering events. Personal protective equipment (PPE) was not used in any observed event. In 442 events (23%), vendors sought assistance from consumers during slaughtering. Slaughtering occurred in plastic jars (1,139 events; 58%), directly on the road (415 events; 21%), in roadside drains (214 events; 11%), and on newspapers (39 events; 2 %). Seventy percent of vendors reported discarding slaughtering waste directly into the environment. The remaining 30% reported storing waste in containers (e.g., plastic drums, buckets, polythene bags, or sacks) sometimes for sale to fisheries or low-income consumers. Observation data showed that waste or dead poultry was discarded on roads, agricultural land, drains, open areas, bushes, ditches, ponds, or rivers. Vendors were also observed feeding slaughtering waste to crows and stray dogs.

#### Hygiene practices and use of personal protective equipment

Nearly 90% of vendors reported being unaware of proper hygiene practices and had never used PPE, such as face masks, aprons, gloves, or head covers. Almost all vendors (38/40) reported not washing their hands after handling poultry. Vendors reported cleaning their tricycle vans or bicycles approximately once per week, while poultry cages were cleaned daily or once every two days. Cages were washed in rivers, ponds, ditches, or at home. Slaughtering equipment was wiped with cloths or rinsed with water; however, most vendors reported cleaning plastic jars used for slaughtering with detergent. All participants reported washing their clothes daily with soap. During observation, vendors repeatedly wiped their hands with cloths (e.g., handkerchiefs) and consumed snacks without washing their hands.

#### Risk perception of avian influenza

Nearly half of the vendors were unfamiliar with the term “bird flu”. Some could describe symptoms observed in poultry. About one-third recognized that poultry diseases could be contagious. Most vendors (78%) reported not knowing how diseases could spread from poultry to other animals or humans, and were unaware of preventive measures. Vendors were unaware of their potential role in disease transmission and/or the importance of safe slaughtering practices.

### Phase 2: Intervention piloting and feedback

During the piloting (Table 1), most vendors (9/10) reported that the killing cone was useful for restraining birds, reducing mess, and improving hygiene during the slaughtering. Vendors noted improved working comfort and environmental cleanliness, including reduced flies and odors. One vendor stated, “The provided killing cone has helped us a lot. Now, blood and waste no longer spread over the road. We can store the slaughtered poultry and waste properly; it’s a real advantage”. Another vendor added, “It looks neat and attracts people. Earlier, customers used to scold us for spreading blood and waste in the environment, but now they appreciate this system”.

Vendors also reported that the informational pamphlet helped reinforce hand hygiene and waste management practices. However, bicycle users reported challenges, including noise, added weight, and reduced balance and speed. One vendor also reported difficulty carrying disinfectant due to a lack of storage space. Although most vendors accepted the cone positively, some noted challenges in maintaining it. One vendor noted, “Blood dries quickly, and we do not always have enough water to wash it properly, even though we try with soap and disinfectant”.

Another vendor stated, “It takes extra time to clean after each slaughter, but it keeps the place neat and stops the bad smell”. Some vendors highlighted trade-offs: One said, “It does not increase or reduce my profit, but it adds extra labor”. Based on this feedback, we made two key refinements to the cone design: (1) the upper opening was widened from 6″ to 8″ to better accommodate larger poultry, and (2) the frame was replaced with a lighter metallic structure to reduce weight and improve stability (Fig. 3). These modifications improved usability and stability before full implementation.

**Fig 3.**
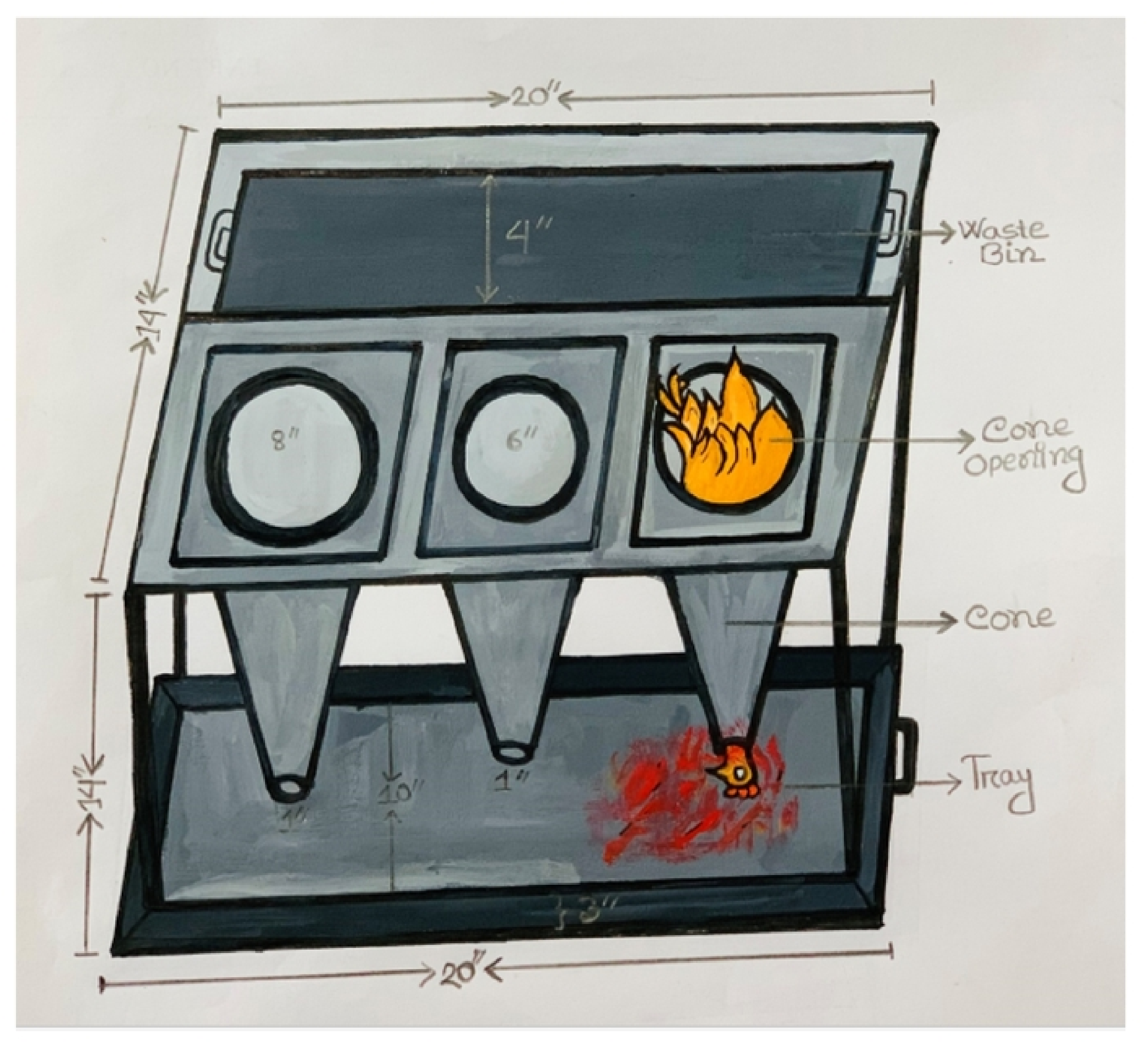
Structural design of the poultry killing cone.

### Phase 3: Implementation and assessment

#### Acceptability of the intervention

Vendors (Table 1) generally perceived the killing cone and waste box as acceptable and useful tools that improved cleanliness and reduced odor. Many reported that the intervention made their practices appear more hygienic to customers. One vendor noted, “After using this cone, customers said the setup looks clean and hygienic. Blood doesn’t scatter on the road, and the environment stays clean.” Another vendor shared a similar experience: “People used to avoid my tricycle because it looked dirty, but now they come and buy more poultry”. Some vendors reported perceived increases in sales. As one explained, “After using this, my daily income increased from 400 to 600 BDT; customers like clean work, so sales have increased”. These were vendor-reported perceptions rather than objectively measured changes in sales.

During implementation, vendors acknowledged that hand sanitizer and waste disinfectants could help prevent disease transmission. Consistent with findings from the pilot phase, vendors using bicycles reported similar challenges with the killing cone; two participants recommended using lighter materials for cones. During participant observation, 294 poultry were slaughtered. Of these, 232 (79%) were slaughtered using a killing cone. Other slaughtering locations included roadside areas (11%), drains (2%), and other locations (7%). Waste was stored in the cone-attached waste boxes in 228 instances (97% of cone uses). Vendors were observed consuming snacks during trading; in 75% of such instances, they cleaned or rubbed their hands using sanitizer before eating. Vendors applied sodium hypochlorite (NaOCl) to waste 39 times, and 75% of vendors disposed of final waste in city corporation dustbins. Vendors did not use PPE at baseline but partially adopted hygiene materials, such as hand sanitizers, during implementation. Among 17 interviewed customers, 88% (15/17) recognized the cone’s role in safer waste disposal, and 53% (9/17) considered it a useful tool overall. These percentages represent proportions of coded responses. Additional perceived benefits included access to freshly slaughtered poultry (29%; 5/17), convenience (24%; 4/17), and reduced exposure to previous slaughter waste (18%; 3/17). Customers also noted environmental cleanliness. One customer stated, “It’s good that the road stays clean, and we are not afraid of any germs”.

## Discussion

The study findings indicate that mobile poultry vending involves multiple practices that may increase opportunities t for exposure to zoonotic pathogens at the human-animal-environment interface. These practices included sourcing poultry from multiple locations, keeping different poultry together in confined cages, slaughtering poultry in open or poorly controlled settings, inadequate cleaning and hand hygiene, unsafe waste disposal, and handling of sick or dead birds (7, 10, 18). These practices, along with limited awareness of disease transmission and preventive measures, highlight important biosecurity gaps among mobile poultry vendors. The intervention package, including the killing cone, waste box, hand sanitizer, disinfectants, and educational pamphlet, was well accepted by vendors and customers and was associated with improved observed slaughtering, waste-management, and hand -hygiene practices during the implementation period. behaviors among. However, because the study did not measure pathogen transmission or environmental contamination, these findings should be interpreted as evidence of improved practices and reduced potential exposure pathways rather than confirmed reductions in infection or transmission.

Since the introduction of HPAI H5N1, 585 poultry outbreaks have been reported across 54 of Bangladesh’s 64 districts. (23). Our study found that vendors sourced multiple poultry species from various locations and housed them together in cages. These cages were often poorly ventilated and lacked regular cleaning or disinfection. A companion study detected influenza A viruses in 17% of poultry samples, including H5 in 4% of chickens and 57% of ducks and H9 in 82% of chickens and 29% of ducks. This companion study also reported an association between door-to-door vending and virus detection (24). Although these findings were from a separate study and were not generated by the present intervention study, they provide relevant context for the poultry-handling practices observed here. Mixing poultry from different sources and species may create opportunities for circulation of avian influenza and other pathogens (18). In addition, slaughtering poultry along roads, in drains, or near water sources may increase opportunities for environmental exposure among people and animals such as dogs and crows. These practices are relevant in the context of recurrent detection of HPAI H5N1in scavenging birds in Bangladesh (25).

Our findings also highlight potential poultry-to-human exposure pathways. Live-bird market trading can provide opportunities for avian influenza virus exposure through the movement and handling of live poultry, slaughtering of sick birds, and inadequate use of protective measures (26). Vendors in our study transported poultry to residential areas and handled and slaughtered birds at customers’ homes and other informal locations (7). During the slaughtering and bleeding, flapping poultry can generate droplets that may contaminate nearby surfaces (27). Daily movement of vendors and poultry, together with potentially contaminated equipment, may also create opportunities for indirect exposure (7). Limited knowledge of avian influenza transmission pathways, combined with the lack of handwashing after handling poultry and before eating, further highlights potential occupational exposure among vendors. Similar exposure pathways have been reported among poultry market workers in Bangladesh (28, 29). International guidelines discourage slaughtering poultry outside designated facilities; however, in low-resource settings, on-site slaughtering may remain common because of consumer preferences for freshly slaughtered poultry, cultural practices, and economic constraints (30, 31). Within this context, the intervention was designed as a practical and low-cost approach to improve hygiene and biosecurity during informal poultry slaughtering. During the pilot phase, most vendors reported that the killing cone helped restrain birds, reduce mess, and improve hygiene. The intervention was further refined based on vendor feedback, including widening the upper opening and replacing the frame with a lighter metallic structure. This co-design process was particularly relevant for mobile vendors because the intervention needed to accommodate different transport modes and operating conditions. During Phase 3, 79% of observed slaughtering events used the killing cone, and waste was stored in the cone-attached waste box in 97% of cone-use events. These observations indicate that the intervention could be incorporated into routine slaughtering practices during the study period. The use of a containment device, with blood collected in a tray and waste stored in a dedicated container, may help reduce direct contact with poultry by-products and opportunities for environmental exposure (32). This approach is consistent with recommendations to strengthen hygiene and biosecurity in poultry-handling environments(33) and with previous research emphasizing improved slaughtering environments, sanitation, and waste management to reduce zoonotic disease risks (34). The intervention also addressed personal hygiene and waste disinfection. At baseline, almost all vendors reported not washing their hands after handling poultry, and no PPE use was observed during the 1,959 slaughtering events. During implementation, vendors used hand sanitizer before eating in 75% of observed eating instances, and sodium hypochlorite was applied to poultry waste on 39 occasions. Evidence from Bangladesh indicates that poultry workers who do not practice hand hygiene may have a higher risk of H5N1 infection (35). Our baseline findings are also consistent with previous studies among poultry workers and small-scale producers, where individuals were observed wiping blood with cloths rather than washing their hands (36). Alcohol-based hand sanitizers containing 62–70% alcohol have been shown to rapidly inactivate HPAI H5N1 viruses (37). These findings support the inclusion of accessible hand-hygiene materials within interventions for informal poultry vendors. However, the observed use of sanitizer and disinfectants should be interpreted as a change in reported or observed practices, rather than evidence that the intervention reduced viral contamination.

Poultry waste is a recognized potential source of pathogens that can affect humans and animals; therefore, appropriate waste management is an important component of environmental hygiene and biosecurity (38, 39). Safe disposal and treatment of poultry waste may also provide environmental benefits through reduced pollution and resource recovery (40). In our study, vendors used sodium hypochlorite to treat waste and disposed of final waste in city corporation dustbins. The use of chlorine-based disinfectants is consistent with evidence on their effectiveness against HPAI H5N1 contamination in poultry environments(41). However, the intervention did not address all potential exposure pathways. In particular, it did not address airborne exposure, the broader occupational risks associated with handling live and sick poultry, or the movement of vendors and poultry between multiple locations. These factors require complementary measures, including appropriate infrastructure, regulatory engagement, risk communication, surveillance, and broader One Health approaches.

Customer feedback further supports the intervention’s practical acceptability. Among the 17 interviewed customers, 15 (88%) recognized the killing cone’s role in safer waste disposal, and 9 (53%) considered it useful overall. Customers also described environmental cleanliness, convenience, and reduced exposure to previous slaughter waste as perceived benefits. These findings suggest that visible improvements in cleanliness may support consumer acceptance of safer slaughtering practices. However, these findings were based on a small purposively selected group of customers and should be interpreted as perceptions rather than evidence of population-level acceptability or changes in purchasing behavior.

The findings have implications for improving biosecurity in informal poultry-vending systems. Mobile vendors operate at the intersection of poultry production, transportation, household consumption, and the environment, yet they often work without access to dedicated slaughtering facilities or formal biosecurity infrastructure. A context-specific intervention that combines a physical containment device with hand hygiene, waste management materials, and simple information may therefore provide a practical option for reducing opportunities for exposure in settings where formal slaughtering facilities are not readily available. However, the intervention should complement rather than replace broader measures addressing poultry movement, surveillance, slaughtering infrastructure, waste disposal systems, and regulatory oversight (42, 43). Further research should assess whether these changes can be sustained over longer periods and whether similar approaches are feasible in other urban and peri-urban settings.

### Limitations

Our study has several limitations. First, it was conducted in only four sub-districts and did not employ random sampling or randomization; therefore, the findings may not be generalizable to all mobile poultry vendors in Bangladesh, and changes observed during Phase 3 cannot be attributed solely to the intervention. Second, a potential Hawthorne effect may have influenced vendor behavior because participants knew they were being observed, although the use of unobtrusive observation during implementation was intended to minimize this effect. Third, some activities may have been missed because observers could not capture all interactions occurring simultaneously. Fourth, the study was conducted over one year, but the number of observed sick or dead poultry may still have varied over time. A 10-year study found no strong seasonal trend in avian influenza among poultry in Bangladesh (44). In addition, the slaughtering and waste-disposal practices observed in our study were consistent with findings from previous studies conducted at different times and locations in Bangladesh(10, 36, 45). Nevertheless, seasonal and temporal variation in vendor practices cannot be completely excluded.

Finally, we did not conduct microbiological or environmental testing for H5N1 or other avian influenza viruses as part of this study. Therefore, our assessment of transmission and exposure risk was based on observed and reported behaviors rather than confirmed viral detection. The companion study detected H5 virus in poultry and influenza A viruses in samples collected from poultry (24), but these findings were generated independently of the present intervention study and should not be interpreted as evidence that the intervention reduced viral contamination or transmission. Similarly, the present study did not include a comparison group or longitudinal follow-up after the intervention period. We therefore cannot determine the extent to which observed changes were sustained or establish the intervention’s effectiveness in reducing infection or environmental contamination.

## Conclusion

This study identified important hygiene and biosecurity gaps among mobile poultry vendors, including the handling of apparently sick poultry, slaughtering in open or poorly controlled locations, inadequate hand hygiene, lack of PPE use, and unsafe disposal of poultry waste. A low-cost intervention package combining a poultry killing cone, waste collection system, hand-hygiene materials, disinfectants, and an informational pamphlet was feasible and generally acceptable to vendors and customers. During the one-month implementation period, vendors adopted the killing cone in most observed slaughtering events, used the associated waste boxes in most cone-use events, and adopted some additional hygiene and waste-management practices.

The findings support the potential use of practical, context-specific interventions to improve biosecurity practices among mobile poultry vendors where formal slaughtering facilities are limited. However, the study did not measure pathogen transmission or environmental contamination and therefore cannot demonstrate a reduction in avian influenza transmission or infection. Future research should include microbiological and environmental assessments, longer follow-up, and broader implementation across different settings to determine the sustainability, effectiveness, and scalability of such interventions. These measures should be integrated with broader One Health strategies addressing surveillance, infrastructure, waste management, poultry movement, and engagement with relevant regulatory authorities.

## Data Availability

Data are available from the data repository committee at icddr,b. The dataset underlying the findings described in the paper cannot be shared publicly due to ethical restrictions related to protecting study participants' privacy and icddr,b’s data access policy (https://www.icddrb.org/policies). icddr,b has a data repository maintains by the research administration. A copy of the complete dataset (anonymized and decoded) of this study will remain at the data repository. Interested researchers may contact Ms. Armana Ahmed, head of research administration, for approval and data access.

## Contributors

MSI and AI conceptualized the study and secured the funding, and developed the methodology. MSI, AI, and MB lead the investigation. Study resources were provided by MSI, AI, MB, KIA, DY, and AMK. Formal analysis was conducted by MB, DY, AMK, and KIA. The original draft was prepared by MB, DY, AMK, and KIA. MSI, MB, and AI provided supervision. Visualization was done by MB, AMK, DY, and MTI. Project administration was managed by MB, MTI, and SSI. All authors contributed to reviewing and editing the manuscript. All authors had full access to the data, critically reviewed the manuscript for important intellectual content, and approved the final version. MSI, AI, and MB had the final responsibility for the decision to submit for publication.

## Data sharing statement

Data are available from the data repository committee at icddr,b. The dataset underlying the findings described in the paper cannot be shared publicly due to ethical restrictions related to protecting study participants’ privacy and icddr,b’s data access policy (https://www.icddrb.org/policies). icddr,b has a data repository maintained by the research administration. A copy of the complete dataset (anonymized and decoded) of this study will remain at the data repository. Interested researchers may contact Ms. Armana Ahmed, head of research administration, for approval and data access.

## Acknowledgments

The authors are grateful to the field team members and study participants from the Dhaka, Mymensingh, and Netrokona districts for their valuable contributions to this project. Sincerely, thank Fiza Feha for preparing the poultry killing cone illustration used in this manuscript. icddr,b gratefully acknowledges the Government of Bangladesh and the Government of Canada for their generous core/unrestricted support. Artificial intelligence tools were used to enhance the readability and English language expression of this manuscript.

## Reference

1. Ahmed SS, Ersbøll AK, Biswas PK, Christensen JP, Hannan AS, Toft N. Ecological determinants of highly pathogenic avian influenza (H5N1) outbreaks in Bangladesh. PLoS One. 2012;7(3):e33938.

2. Haque M, Giasuddin M, Chowdhury E, Islam M. Molecular evolution of H5N1 highly pathogenic avian influenza viruses in Bangladesh between 2007 and 2012. Avian Pathology. 2014;43(2):183–94.

3. Brooks WA, Alamgir AS, Sultana R, Islam MS, Rahman M, Fry AM, et al. Avian influenza virus A (H5N1), detected through routine surveillance, in child, Bangladesh. Emerg Infect Dis. 2009;15(8):1311– 3.

4. Chakraborty A, Rahman M, Hossain MJ, Khan SU, Haider MS, Sultana R, et al. Mild Respiratory Illness Among Young Children Caused by Highly Pathogenic Avian Influenza A (H5N1) Virus Infection in Dhaka, Bangladesh, 2011. The Journal of Infectious Diseases. 2017;216(suppl_4):S520–S8.

5. Wang Z, Li H, Li Y, Wu Z, Ai H, Zhang M, et al. Mixed selling of different poultry species facilitates emergence of public-health-threating avian influenza viruses. Emerging Microbes & Infections. 2023;12(1):2214255.

6. Pinotti F, Kohnle L, Lourenço J, Gupta S, Hoque MA, Mahmud R, et al. Modelling the transmission dynamics of H9N2 avian influenza viruses in a live bird market. Nature Communications. 2024;15(1):3494.

7. Van Kerkhove MD, Vong S, Guitian J, Holl D, Mangtani P, San S, et al. Poultry movement networks in Cambodia: implications for surveillance and control of highly pathogenic avian influenza (HPAI/H5N1). Vaccine. 2009;27(45):6345–52.

8. Moyen N, Hoque MA, Mahmud R, Hasan M, Sarkar S, Biswas PK, et al. Avian influenza transmission risk along live poultry trading networks in Bangladesh. Scientific Reports. 2021;11(1):19962.

9. Rimmelzwaan GF, van Riel D, Baars M, Bestebroer TM, van Amerongen G, Fouchier RA, et al. Influenza A virus (H5N1) infection in cats causes systemic disease with potential novel routes of virus spread within and between hosts. The American journal of pathology. 2006;168(1):176–83.

10. Rimi NA, Sultana R, Ishtiak-Ahmed K, Khan SU, Sharker M, Zaman RU, et al. Poultry slaughtering practices in rural communities of Bangladesh and risk of avian influenza transmission: a qualitative study. EcoHealth. 2014;11(1):83–93.

11. Organization WH. Critically important antimicrobials for human medicine: 6th revision. Geneva; 2019.

12. Nations FaAOotU. The State of Food and Agriculture 2020: Overcoming water challenges in agriculture. Rome; 2020.

13. Olsen Br, Munster VJ, Wallensten A, Waldenström J, Osterhaus AD, Fouchier RA. Global patterns of influenza A virus in wild birds. science. 2006;312(5772):384–8.

14. Meseko C, Oluwayelu D. Avian influenza. Transboundary Animal Diseases in Sahelian Africa and Connected Regions: Springer; 2019. p. 345–74.

15. Hay AJ, McCauley JW. The WHO global influenza surveillance and response system (GISRS)—a future perspective. Influenza and other respiratory viruses. 2018;12(5):551–7.

16. Pinto J, Von Dobschuetz S, Kondolas G, Larfaoui F, Hamilton-West C. FAO EMPRES-i: an integrated tool for early warning and animal disease surveillance. Challenges of Animal Health Information Systems and Surveillance for Animal Diseases and Zoonoses. 2011:115.

17. Szablewski CM, Iwamoto C, Olsen SJ, Greene CM, Duca LM, Davis CT, et al. Reported global avian influenza detections among humans and animals during 2013-2022: comprehensive review and analysis of available surveillance data. JMIR Public Health and Surveillance. 2023;9(1):e46383.

18. Chowdhury S, Azziz-Baumgartner E, Kile JC, Hoque MA, Rahman MZ, Hossain ME, et al. Association of Biosecurity and Hygiene Practices with Environmental Contamination with Influenza A Viruses in Live Bird Markets, Bangladesh. Emerg Infect Dis. 2020;26(9):2087–96.

19. Mienur Meher M, Afrin M, Bayazid AA, Parajuli N, Rahman MS, Islam MS, et al. Elucidating the perception, awareness and biosecurity practices among the live poultry/poultry-meat sellers to mitigate the risk of zoonotic salmonellosis. bioRxiv. 2025:2025.06. 17.660267.

20. Ekong PS, Ducheyne E, Carpenter TE, Owolodun OA, Oladokun AT, Lombin LH, et al. Spatio-temporal epidemiology of highly pathogenic avian influenza (H5N1) outbreaks in Nigeria, 2006–2008. Preventive veterinary medicine. 2012;103(2-3):170–7.

21. Spickler AR, Trampel DW, Roth JA. The onset of virus shedding and clinical signs in chickens infected with high-pathogenicity and low-pathogenicity avian influenza viruses. Avian Pathology. 2008;37(6):555–77.

22. Benton JS, French DP. Untapped Potential of Unobtrusive Observation for Studying Health Behaviors. JMIR Public Health Surveill. 2024;10:e46638.

23. Islam A, Ara T, Amin E, Islam S, Sayeed MA, Shirin T, et al. Epidemiology and Evolutionary Dynamics of High Pathogenicity Avian Influenza (HPAI) H5N1 in Bangladesh. 2023;2023(1):8499018.

24. Ausraful Islam MAR, Rahul Deb Sarker, Mohammed Ziaur Rahman, Mohammad Enayet Hossain, M. Saiful Islam. Mobile poultry vendors in Bangladesh: practices and movement patterns. Unpublished Result. 2026;In submission.

25. Islam A, Islam S, Hossain ME, Samad MA, Billah MM, Hassan MM, et al. One Health Investigation of House Crow (Corvus splendens) Mortality Event Linked to the Potential Circulation of H5N1 Virus at Live Bird Markets in Northwestern Bangladesh. International Journal of Infectious Diseases. 2022;116:S112.

26. Fournié G, Guitian J, Desvaux S, Cuong VC, Dung DH, Pfeiffer DU, et al. Interventions for avian influenza A (H5N1) risk management in live bird market networks. Proceedings of the National Academy of Sciences. 2013;110(22):9177–82.

27. Wang X, Wang Q, Cheng W, Yu Z, Ling F, Mao H, et al. Risk factors for avian influenza virus contamination of live poultry markets in Zhejiang, China during the 2015–2016 human influenza season. 2017;7(1):42722.

28. Hassan MZ, Sturm-Ramirez K, Islam MS, Afreen S, Rahman MZ, Kafi MAH, et al. Interpretation of molecular detection of avian influenza A virus in respiratory specimens collected from live bird market workers in Dhaka, Bangladesh: infection or contamination? International journal of infectious diseases : IJID : official publication of the International Society for Infectious Diseases. 2023;136:22–8.

29. Nasreen S, Khan SU, Luby SP, Gurley ES, Abedin J, Zaman RU, et al. Highly pathogenic Avian Influenza A(H5N1) virus infection among workers at live bird markets, Bangladesh, 2009-2010. Emerging infectious diseases. 2015;21(4):629–37.

30. Sultana R, Nahar N, Rimi NA, Azad S, Islam MS, Gurley ES, et al. Backyard poultry raising in Bangladesh: a valued resource for the villagers and a setting for zoonotic transmission of avian influenza. A qualitative study. Rural and remote health. 2012;12:1927.

31. Nielsen SS, Alvarez J, Bicout DJ, Calistri P, Depner K, Drewe JA, et al. Slaughter of animals: poultry. EFSA journal European Food Safety Authority. 2019;17(11):e05849.

32. Bertran K, Clark A, Swayne DE. Mitigation strategies to reduce the generation and transmission of airborne highly pathogenic avian influenza virus particles during processing of infected poultry. International journal of hygiene and environmental health. 2018;221(6):893–900.

33. Organization WH. Public health interventions for prevention and control of avian influenza. 2006.

34. Rimi NA, Hassan MZ, Chowdhury S, Rahman M, Sultana R, Biswas PK, et al. A Decade of Avian Influenza in Bangladesh: Where Are We Now? Tropical Medicine and Infectious Disease. 2019;4(3):119.

35. Nasreen S, Khan SU, Luby SP, Gurley ES, Abedin J, Zaman RU, et al. Highly pathogenic avian influenza A (H5N1) virus infection among workers at live bird markets, Bangladesh, 2009–2010. Emerging infectious diseases. 2015;21(4):629.

36. Hasan SM, Sturm-Ramirez K, Kamal A-HM, Islam MA, Rahman M, Kile JC, et al. Quail Rearing Practices and Potential for Avian Influenza Virus Transmission, Bangladesh. EcoHealth. 2023;20(2):167– 77.

37. Bondi C, Arbogast J, Macinga D, Lambkin-Williams R, Moane E. Virucidal performance of various professional hand hygiene products against avian influenza A H5N1. American Journal of Infection Control. 2007;35(5):E34–E5.

38. Mozhiarasi V, Natarajan TS. Slaughterhouse and poultry wastes: management practices, feedstocks for renewable energy production, and recovery of value added products. Biomass Conversion and Biorefinery. 2022.

39. Muduli S, Champati A, Popalghat HK, Patel P, Sneha K. Poultry waste management: An approach for sustainable development. Int J Adv Sci Res. 2019;4(1):8–14.

40. Rahman MM, Hassan A, Hossain I, Jahangir MMR, Chowdhury EH, Parvin R. Current state of poultry waste management practices in Bangladesh, environmental concerns, and future recommendations. Journal of Advanced Veterinary and Animal Research. 2022;9(3):490.

41. Rohaim MA, El-Naggar RF, Gamal AM, Ismael E, Hamoud MM, Moubarak ST, et al. Efficacy of disinfectants against egyptian H5N1 avian influenza virus. Hosts and Viruses. 2015;2(5):80.

42. Islam A, Rahman MZ, Hassan MM, Epstein JH, Klaassen M. Farm biosecurity practices affecting avian influenza virus circulation in commercial chicken farms in Bangladesh. One Health. 2024;18:100681.

43. Ahmed HA, Mohamed ME, Erfan AM, Awadallah MA, Abdelkarim L. Investigating the biosecurity measures’ applications in poultry farms and its relationship with the occurence of avian influenza. 2021.

44. Berry I, Rahman M, Flora MS, Shirin T, Alamgir A, Khan MH, et al. Seasonality of influenza and coseasonality with avian influenza in Bangladesh, 2010–19: a retrospective, time-series analysis. The Lancet Global Health. 2022;10(8):e1150–e8.

45. Berry I, Rahman M, Flora MS, Greer AL, Morris SK, Khan IA, et al. Frequency and patterns of exposure to live poultry and the potential risk of avian influenza transmission to humans in urban Bangladesh. Scientific reports. 2021;11(1):21880.

